## Supplement for "COVID-19 vaccination in Sindh Province, Pakistan: a modelling study of health impact and cost-effectiveness"

#### 1. Model Details

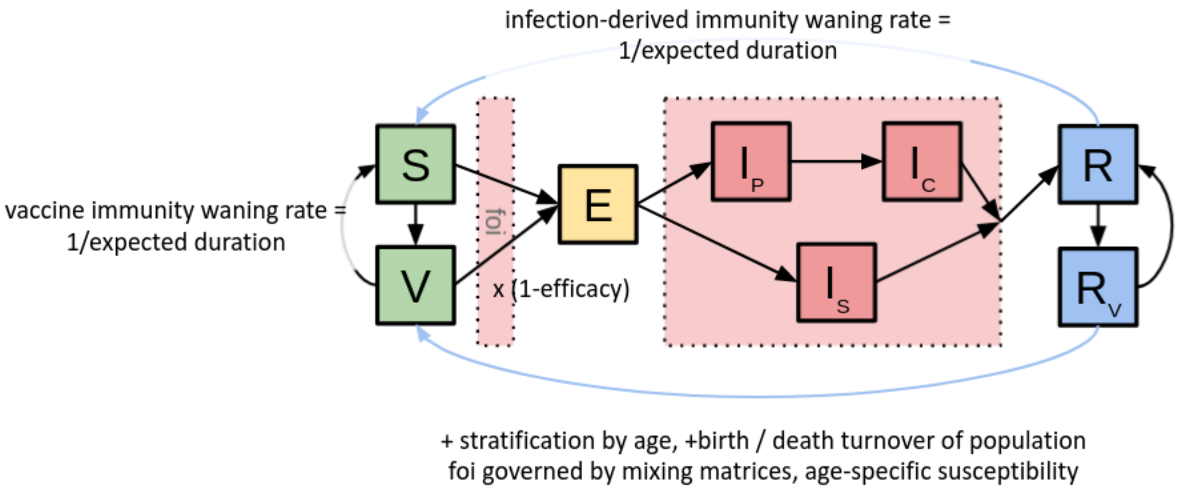


***Figure S1: Model diagram.***

We used a previously published compartmental model (1–3) tailored to the population of Sindh using population data from WorldPop (4) and assumed baseline population contact rates from previously estimated national patterns for Pakistan (5).

The model compartments are an extended SEIRS+V (*S*usceptible, *E*xposed, *I*nfectious with multiple sub-compartments, *R*ecovered and *V*accinated, both returning to *S*usceptible) system with births, deaths, and age structure. We represent the Infectious state with two paths: some individuals remain subclinical, but still transmit, while other individuals undergo a pre-symptomatic state then a symptomatic state. We assume that the subclinical path is half as infectious as the symptomatic path. For all compartments other than *R*ecovered and *V*accinated, we use event-time distributions derived from global observations (Main text Table 1). For *R*ecovered and *V*accinated, we consider multiple characteristic protection durations, given the uncertainty in these durations. We do not assume specific reasons for returning to *S*usceptible; these could represent e.g. antibody waning, emergence of new variants, or some mixture of mechanisms. For the *R*ecovered compartment, we assume perfect protection; we address the Vaccinated compartment protection along with the vaccination programme details in the “Vaccine Programme" section.

We assume that age groups have different susceptibility to infection and, upon infection, different probabilities of following the subclinical versus symptomatic paths. These parameters are from an analysis using data from six countries (1). For disease severity outcomes, we assume the same age-specific hospitalisation, ICU, and death risk as used in previous work (2).

We assumed that contact patterns changed over the course of the epidemic (with corresponding changes in the time-varying effective reproduction number, $R_t$), and estimated these changes using Google Community Mobility indicators (6) for Sindh and school closures as reflected in the Oxford Coronavirus Government Response Tracker (7). We represent contact matrices with four components, corresponding to contacts associated with home, work, school, and miscellaneous "other" settings. We assumed home contacts scale linearly with the Google Mobility indicator for residential visits, work contacts scale linearly with the indicator for workplace visits, and school contacts are zero when schools are closed but unchanged otherwise.

For "other" contacts, we aggregate several Google Mobility indicators ($g_{groc}$, grocery and pharmacy; $g_{ret}$, retail and recreation; $g_{tra}$, transit; $g_{par}$, parks), and then scale linearly with the following weighted aggregation: $0.3(g_{groc} + g_{ret} + g_{tra}) + 0.1g_{par}$. For all indicators, $g_* = 1$ corresponds to the average measured during the first two weeks of February 2020. We assume that $g_* = 1$ corresponds to the contact matrices estimated for Pakistan in (5), $g_* = 0$ corresponds to no contacts, and linear scaling for other values. See SI Figure 1 for a model diagram and SI Figure 2 for population demography and contact patterns.

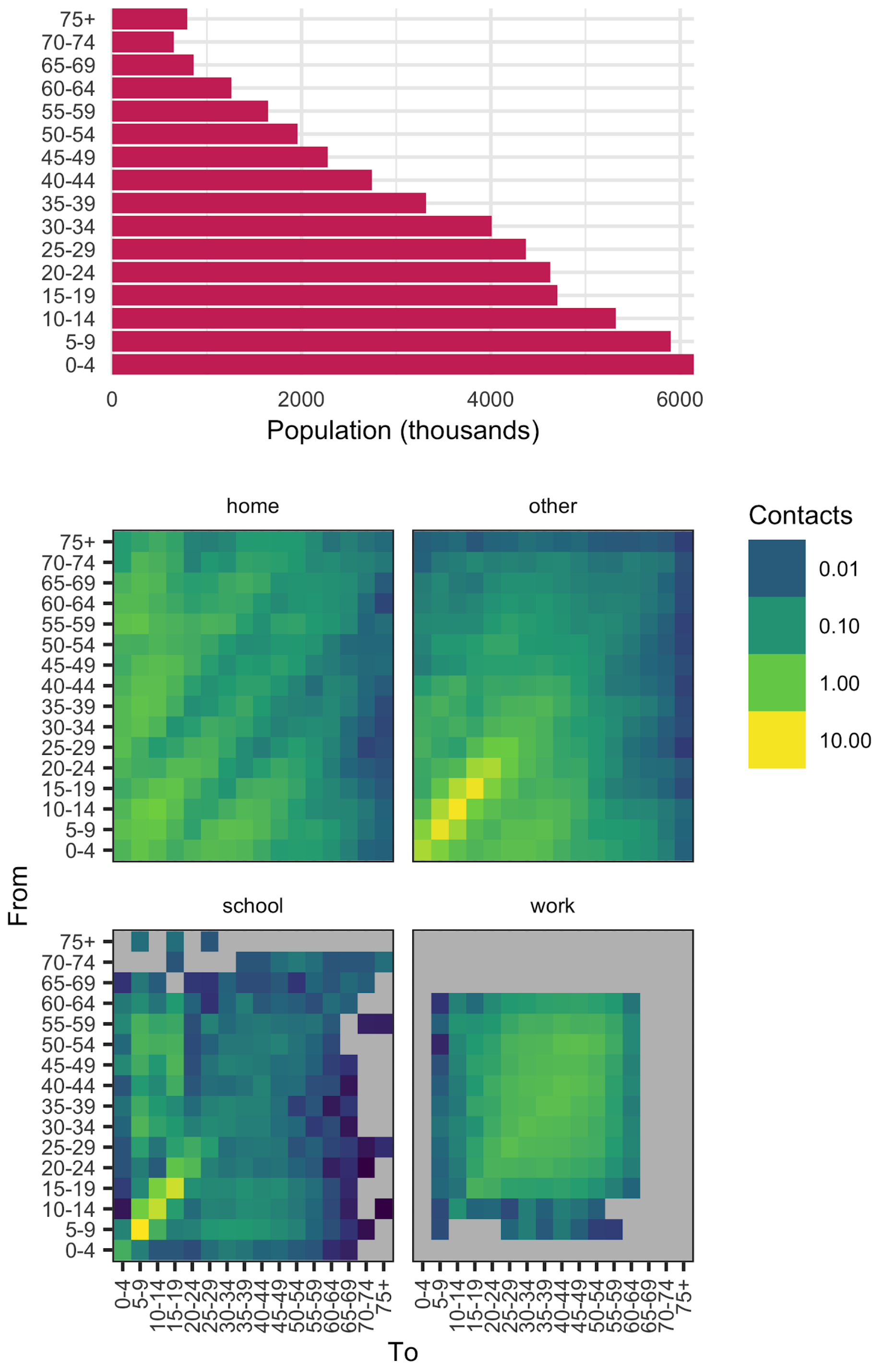


***Figure S2: Demographics in the modelled population.***
Demography of Singh province, Pakistan.

#### 2. Model fitting to COVID-19 cases and deaths in Sindh

For introduction date, we assume that 10 SARS-CoV-2 infections are seeded in Sindh province on day $t_0$, adopting a uniform prior probability for $t_0$ over the first 120 days of 2020; this is meant to approximately capture when sustained community transmission of

SARS-CoV-2 began rather than represent a precise schedule of infection importations, since continued importation will eventually become negligible relative to exponentially growing local transmission. For transmission, we adopted a vague prior for the basic reproduction number, following a normal distribution centered on $R_0$ = 2.4 with standard deviation 1.2.

The ascertainment proportion for both cases and deaths is parameterised using a starting value (i.e., the ascertainment proportion on 1 January 2020), an end value (the ascertainment proportion on 6 September 2020), and two additional parameters that determine the shape of the ascertainment rate between these two points. We model time-varying ascertainment rates for both cases and deaths as

$$asc(t; t_{max}, a_0, a_1, s_0, s_1) = a_0 + (a_1 - a_0)(lo(s_0 + (t/t_{max})(s_1 - s_0)) - lo(s_0))/(lo(s_1) - lo(s_0))$$

where

$$lo(x) = (exp\, x)/(1 + exp\, x).$$

Above, $asc(t; t_{max}, a_0, a_1, s_0, s_1)$ is a logistic-shaped curve parameterized to be a smooth S-shaped function of $t$ from 0 to $t_{max}$, which goes from $a_0$ at $t = 0$ to $a_1$ at $t = t_{max}$, with an inflection point at $t = -s_0 t_{max}/(-s_0 + s_1)$ if $s_0 < 0$ and $s_1 > 0$. As prior distributions for the parameters governing time-varying case and death ascertainment rates, we assume
$a_0^{cases}, a_0^{deaths} \sim uniform(0, 1)$,
$log(a_1^{cases}/a_0^{cases}), log(a_1^{deaths}/a_0^{deaths}) \sim normal(0, 0.5)$, and
$-s_0^{cases}, -s_0^{deaths}, s_1^{cases}, s_1^{deaths} \sim exponential(1)$.

We assume that observed cases and deaths follow a normal distribution centered on model-predicted reported cases and deaths with standard deviations $\sigma_{cases} \sim halfnormal(0, 50)$ and $\sigma_{deaths} \sim halfnormal(0, 5)$. It is more conventional to model epidemiological incidence data using a Poisson or negative binomial distribution, but we found that a normal distribution resulted in a better fit, possibly because it was less sensitive to noisy data. Specifically, the likelihood of the model fit was

$$L = \prod_j \phi(c_j | \mu = C(t_j)\, asc(t_j; t_{max}, a_0^{cases}, a_1^{cases}, s_0^{cases}, s_1^{cases}), \sigma = \sigma_{cases})$$
$$\times \prod_k \phi(d_k | \mu = D(t_k)\, asc(t_k; t_{max}, a_0^{deaths}, a_1^{deaths}, s_0^{deaths}, s_1^{deaths}), \sigma = \sigma_{deaths}),$$

where $\phi(x|\mu, \sigma)$ is the normal probability density function, $c_j$ is the observed number of reported cases on day $t_j$, $C(t_j)$ is the model-predicted number of true case onsets on day

$t_j$, $d_k$ is the observed number of reported deaths on day $t_k$, and $D(t_k)$ is the model-predicted number of true COVID-19 deaths on day $t_k$.

#### 3. Alternative Fitting Assumptions

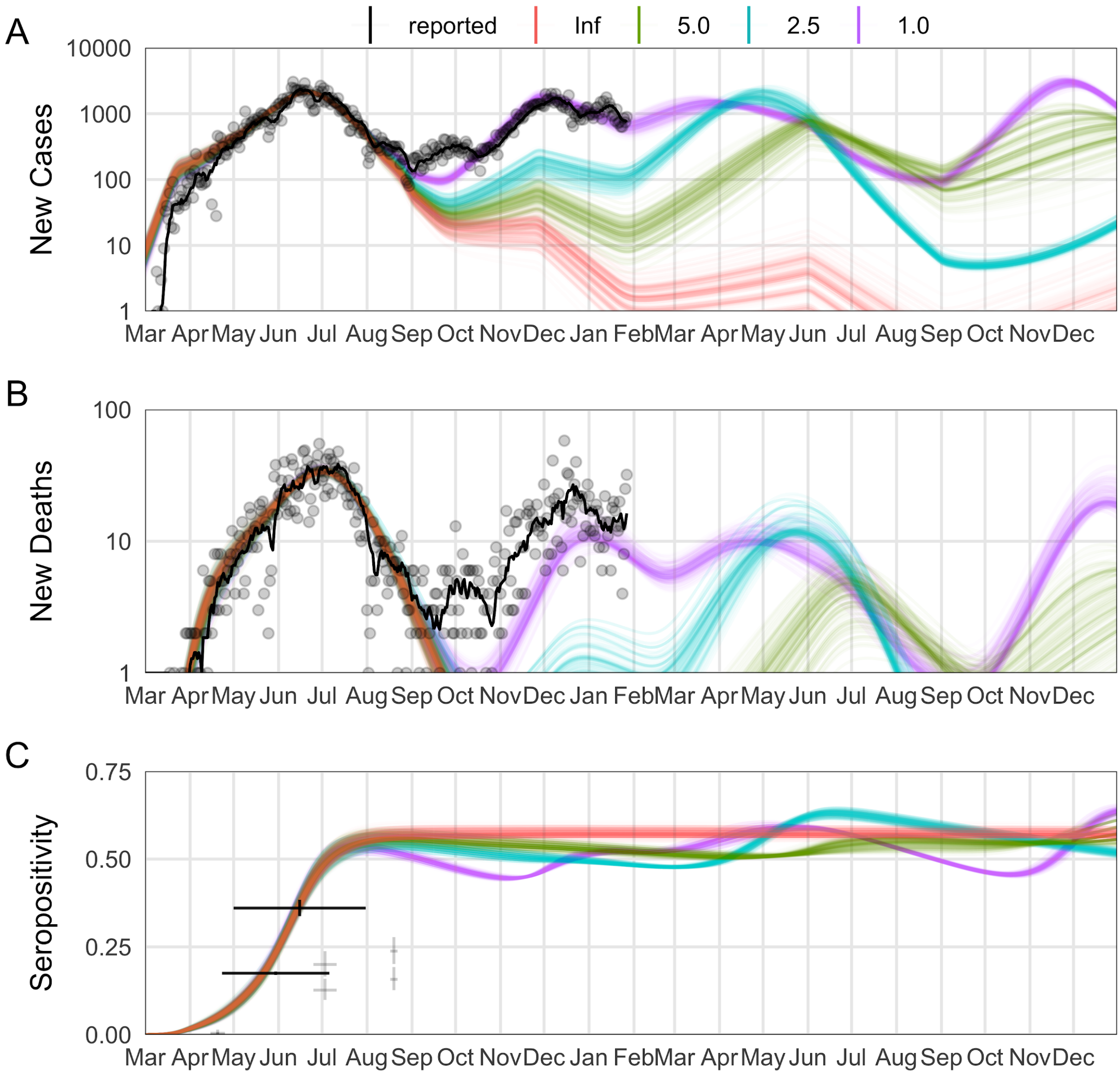


***Figure S3: Alternative Infection-Induced Protection Durations.***
These panels show the various durations assumed for infection-induced immunity. As can be seen across all assumptions, they can produce comparable fits to the data up to September 2020, but as they extend into 2021, the different assumptions diverge in both timing and magnitude.

### 4. Additional Health Outcomes

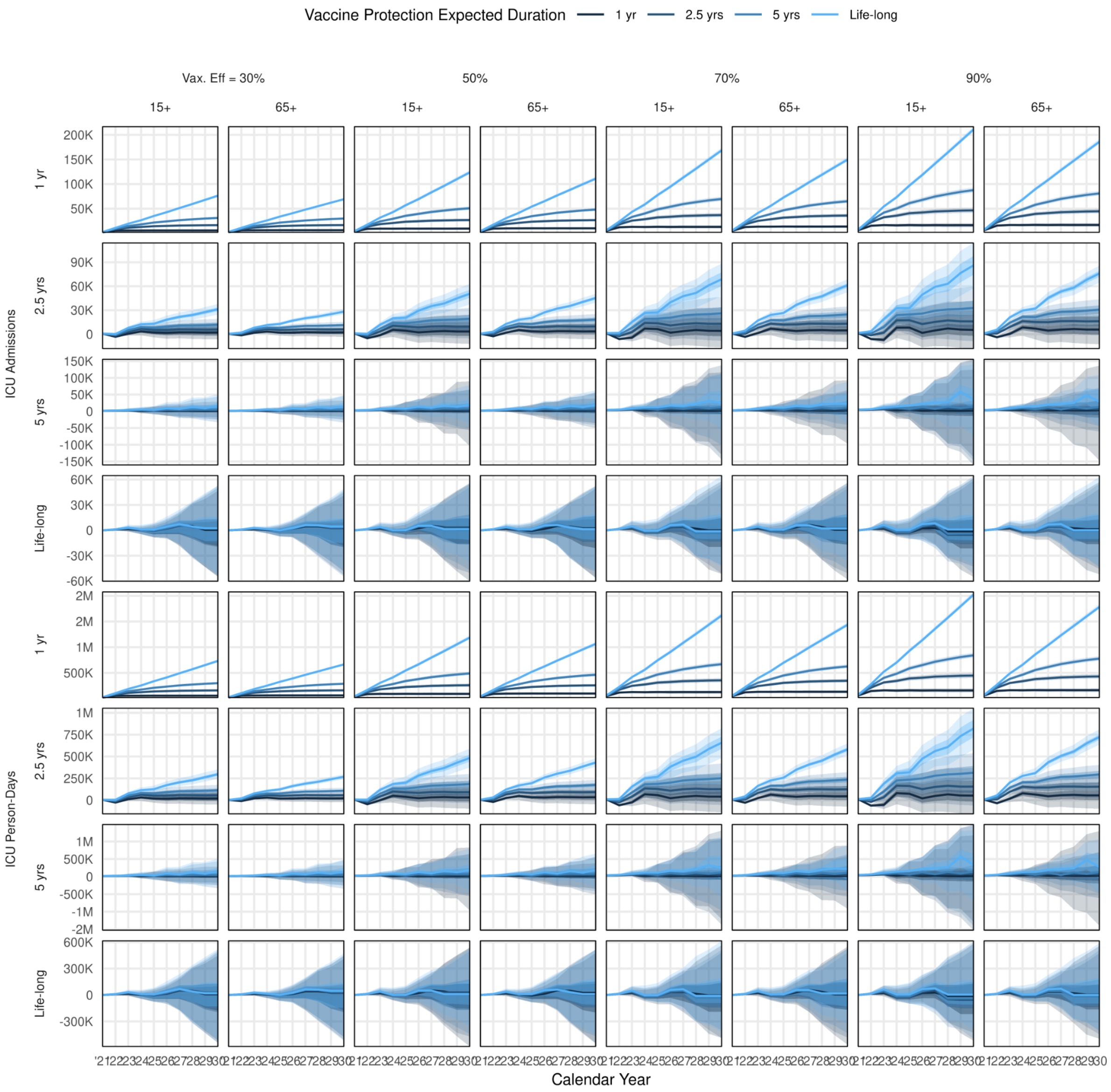


***Figure S4: Trends in ICU admissions and person-days.***
Similar to Figure 3 in the main text, we also project hospital admissions and occupancy, for both intensive / critical care (ICU, this figure) and general hospital care (Ward, Supplement Figure 5). The trends for hospitalisation are very similar to those in cases and deaths.

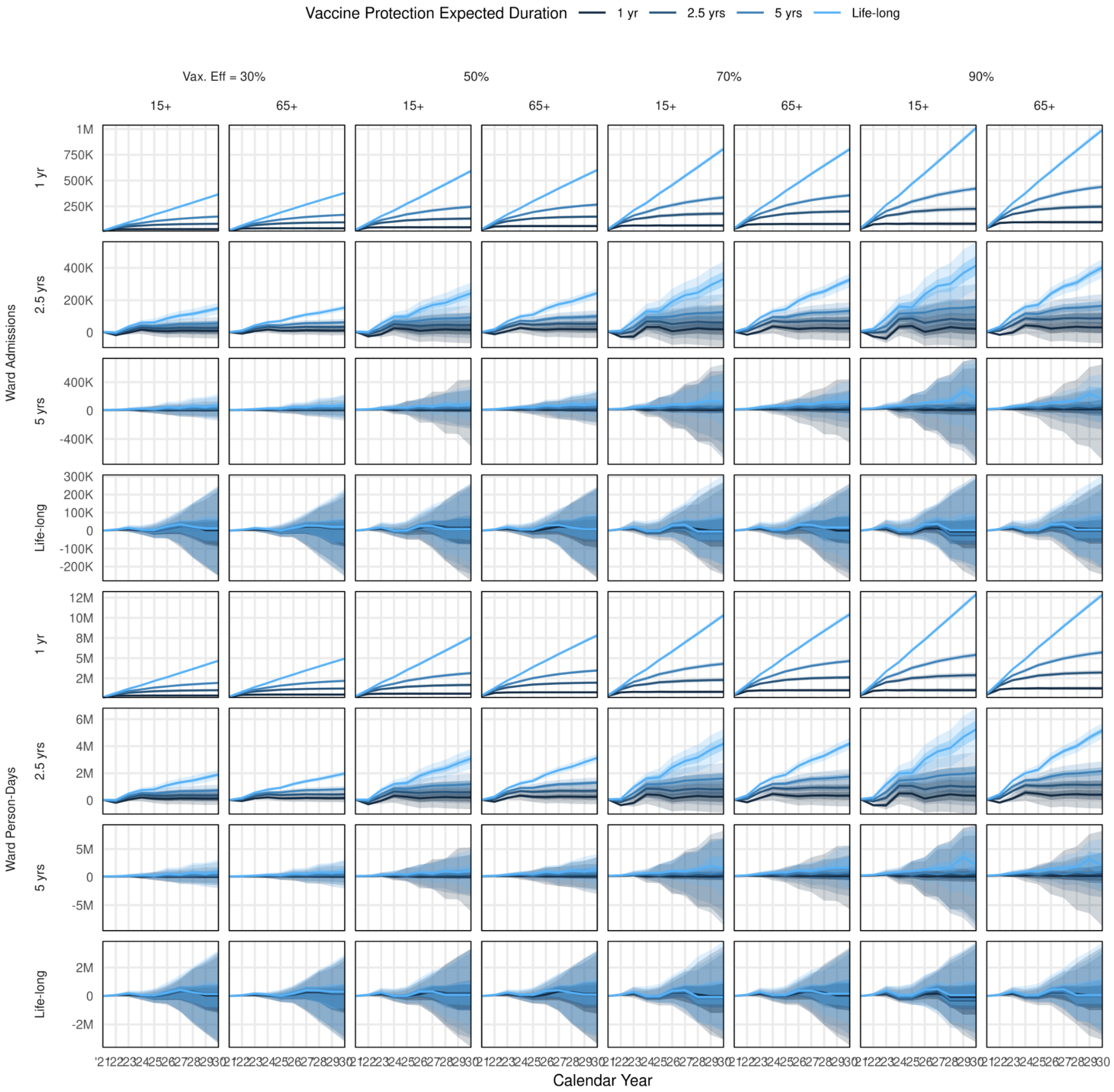


***Figure S5: Trends in General Ward Admissions and person-days.***

#### 5. Number of vaccine doses available

In Phase 1 of its vaccine allocation, COVAX aims to make available vaccine doses according to participating countries' populations. An initial tranche of vaccine doses to cover 3% of the population of these countries is intended to be made available, expanding to 20% by the end of Phase 1 (8).

A projected timeline over which the doses will be available has been provided by WHO SAGE for mathematical modellers (9). We used this timeline (excluding direct country procurement) in order to project the number of doses available in Sindh. Given that Sindh has a population of 48 million, out of a global population of 7.8 billion but with only 70% in countries participating in COVAX (10), we assumed that 0.88% of COVAX doses will be allocated to Sindh.

Vaccines were not available in 2020, so we ignored the first row. In addition, we pushed the start of vaccination by 3 months compared to the WHO SAGE timeline due to delays to the start of COVAX allocations and likely time it will take for the vaccines to reach Sindh.

Given the doses available, we assume these are distributed every day of a quarter. For two-dose courses, the daily doses available are therefore the total doses available, times 90% (for 15% wastage), divided by 182. This comes out to 4110 per day, which we pessimistically round to 4000 per day.

***Table S1. Timeline for COVAX dose availability.***
Q4 2020 is removed because doses were not available at that time.

| Timeline (WHO SAGE) | Timeline (us) | Doses available (total) | Doses available (Sindh) |
|---|---|---|---|
| ~~Q4 2020~~ | ~~Q2 2021~~ | ~~100 million~~ | ~~880,000~~ |
| Q1 2021 | Q2 2021 | 100 million | 880,000 |
| Q2 2021 | Q3 2021 | 400 million | 3,500,000 |
| Q3 2021 | Q4 2021 | 600 million | 5,270,000 |
| Q4 2021 | Q1 2022 | 800 million | 7,030,000 |

### 6. Costs of COVID-19 vaccine introduction

***Vaccine and Immunisation Supplies***
We assumed a procurement price of the vaccine of $3 per dose (10), with an additional 10% added for freight costs. We also account for the costs of an AD syringe and safety box. We assumed a 15%, the wastage typically experienced during campaigns (11).

***Cold chain***
Cold chain costs were divided between national level and service level. National level costs were obtained from (11). Costs were calculated by estimating the additional cold chain equipment needed at national, regional and district levels, including walk-in cold rooms, ice-lined refrigerators, solar direct drive refrigerators and freezers for ice pack preparations.

Pakistan was scheduled to undergo a national- and provincial-level expansion of vaccine cold storage facilities in 2017-18. Existing cold storage space was estimated by adding the required 2017 expansion volume to the available volume reported by the latest Effective Vaccine Management report (12). Additional cold chain costs at the service level were estimated by assuming a packed volume per dose equivalent to that of a comparable vaccine (4.8cm$^3$ for 1-dose vials, as for tetanus toxoid vaccines). We estimated the volume of the COVID-19 vaccine relative to other vaccines in the immunisation programme in order to allocate a COVID-19 vaccine-specific proportion of equipment and electricity costs. Facility-level cold storage equipment prices were obtained from the UNICEF Supply Divisions price lists (13). We obtained electricity consumption data for each type of equipment from product information sheets. We multiplied the total number of kilowatt hours required by the price per kilowatt hours in Pakistan (14).

***Human resources***
We assumed two types of healthcare workers would perform vaccinations: nurses and vaccinators. Monthly salaries for both types were sourced from federal pay scales and a cost

per minute of work was calculated (2.83 PKR for nurses and 2.20 for vaccinators). We assumed that one health care worker would require 20 minutes to carry out a vaccination and all other relevant processes per person vaccinated. In addition, we assumed the health care worker would require 7 additional minutes devoted to transport per person vaccinated. Time use data were obtained from cost estimates of the tetanus toxoid vaccine, estimated for the Disease Control Priorities project in Pakistan (15,16).

***Transport***

We divided transport costs between those required to get vaccines to the facility (facility-based), and those required at the facility level in order to deliver vaccines in campaigns (campaign-based). We obtained facility-based costs from UNICEF (11); they reviewed available costing studies in low- and middle-income countries, finding a median value of $0.04, which accounts for storage and transport (including per diems related to these activities).

We estimated campaign-based costs by calculating fuel and vehicle maintenance costs per campaign trip. We estimated the size of health facility catchment areas at 66 $km^2$ by dividing the total number of health facilities in the country by the total area of the country. We calculated the radius of the catchment area ($A=\pi r^2$) to be 4.59km and assumed a daily trip of 2x radius. We assumed that campaigns would take place during all 261 working days per year across all health facilities in Sindh Province, estimated at 1931 (17). We assumed the fuel costs to be $0.65 per litre, and a fuel efficiency of 15 km per litre (18). We assumed a cost of vehicle maintenance per km of $0.09 obtained from WHO-CHOICE (19).

***Social mobilisation***

We obtained the costs of social mobilisation per dose from budgeting data for a poliovirus detection and interruption campaign in Pakistan in 2019. Data were obtained from the Polio Global Eradication Initiative (20).

***Health system mark-up***

We applied a 31% mark-up to our delivery costs (i.e. all above mentioned costs excluding vaccine and immunisation supplies) in order to account for the following indirect but related activities: planning and coordination, training, pharmacovigilance, vaccination certificates, personal protective equipment, hand hygiene, waste management and technical assistance. This mark-up was derived by obtaining the costs of these activities relative to all delivery costs from UNICEF (11).

### 7. Costs of COVID-19 clinical management

The main parameters used in the estimates of health resources and costing of COVID-19 response and case management are summarised below. A set of COVID-19 diagnosis and treatment interventions was identified following WHO guidance shown in Table S1. A corresponding list of unit costs was thus generated, to be brought together with the epidemiological model for estimating financial resource needs. More on the costs of COVID-19 clinical management can be found in Torres-Rueda et al. 2020 (21).

***Table S2. Intervention description and unit costs***

| Activity | Activity costs |
| --- | --- |
| Case finding, contact tracing and management: Contact tracing | Per person contacted |
| Case finding, contact tracing and management: Quarantine of contacts | Per person quarantined |
| Screening and diagnosis | Per person screened and tested |
| Case Management: Home-based care | Per person requiring home-based care |
| Case Management: Hospital-based (severe case) | Per day of hospitalisation (severe case) |
| Case Management: Hospital-based (critical case) | Per day of hospitalisation (critical case) |
| Case Management: Death | Per COVID-related death |

For each of the activities mentioned above, we used ingredients-based costing to identify a series of inputs (e.g. junior-level government worker day). For each input we estimated a number of units (e.g. three days of work) and a price. The costs of each input were identified using a range of sources, according to availability of recent primary cost data and appropriateness of cost estimates to the COVID pattern of care. Quantities and prices are presented in Table S3.

***Table S3. Quantities and price of inputs and unit costs per activity***

| Inputs | Number of Units per Input | Price per Input (USD) |
| --- | --- | --- |
| **Case finding, contact tracing and management: Contact tracing** | | |
| Working day (junior level govt) | 0.1 | 13.07 |
| Contact tracing household visit | 0.33 | 3.02 |
| Contact tracing phone call | 0.67 | 0.34 |
| **Unit Cost:** | | ***2.54*** |
| **Case finding, contact tracing and management: Quarantine of contacts** | | |
| Working day (health care workers) | 0.1 | 10.43 |
| Working day (junior level govt) | 0.1 | 13.07 |
| **Unit Cost:** | | ***2.35*** |
| **Screening and diagnosis** | | |
| Ambulance trip | 0.0001 | 9.51 |
| Isolation pod/ diagnostic visit | 2 | 0.49 |
| Outpatient visit oral history | 1 | 0.47 |
| Outpatient visit physical exam | 1 | 0.47 |
| Outpatient visit specimen collection | 1 | 1.09 |
| COVID19 test (PCR) | 1 | 23.98 |
| **Unit Cost:** | | ***26.98*** |

| | | |
|---|---|---|
| **Case Management: Home-based care** | | |
| Home-based care bed-day | 5 | 0.61 |
| Community-based care via GP | 2 | 4.71 |
| **Unit Cost:** | | ***12.45*** |
| **Case Management: Hospital-based (severe case)** | | |
| Inpatient ward bed-day (severe) | 1 | 31.54 |
| *Diagnostics* | | |
| Pulse oximetry | 0.125 | 0.00 |
| Chest X-ray | 0.125 | 2.79 |
| Full blood count | 0.125 | 2.29 |
| Blood urea and electrolyte test | 0.125 | 2.53 |
| C-reactive protein test | 0.125 | 0.32 |
| HIV test | 0.125 | 3.87 |
| COVID19 test (PCR) | 0 | 23.98 |
| Malaria test | 0.125 | 0.19 |
| Haemoglobin test | 0.125 | 2.29 |
| **Unit Cost:** | | ***33.32*** |
| **Case Management: Hospital-based (critical case)** | | |
| Inpatient ward bed-day (critical) | 0.33 | 32.29 |
| ITU bed-day | 0.67 | 101.99 |
| Additional resourcing per COVID-related complication | | |
| Acute respiratory distress syndrome (ARDS) | 0.47 | 22.46 |
| Acute kidney injury days | 0.04 | 10.60 |
| Acute cardiac injury days | 0.06 | 46.25 |
| Liver dysfunction days | 0.06 | 89.32 |
| Pneumothorax days | 0.01 | 6.77 |
| Hospital-acquired pneumonia days | 0.05 | 18.85 |
| Bacteraemia days | 0.01 | 32.55 |
| Urinary tract infection days | 0.01 | 9.03 |
| Septic shock days | 0.05 | 0.67 |
| *Diagnostics* | | |
| Pulse oximetry | 10 | 0.00 |
| Chest X-ray | 10 | 2.79 |
| Full blood count | 10 | 2.29 |
| Blood urea and electrolyte test | 10 | 2.53 |
| C-reactive protein test | 10 | 0.32 |

| | | |
|---|---|---|
| Venous blood gas test | 10 | 4.23 |
| HIV test | 0.1 | 3.87 |
| COVID19 test (PCR) | 0 | 23.98 |
| Malaria test | 0.1 | 0.19 |
| Haemoglobin test | 0.1 | 2.29 |
| **Unit Cost:** | | **221.18** |
| **Case Management: Death** | | |
| Body Bag | 1 | 64.52 |
| **Unit Cost:** | | **64.52** |

***Clinical Management***

The number of days per patient in general ward and in ICU was set at 8 and 10 respectively, and was set to match the assumptions in the epidemiological model (22,23). Following expert clinician advice we assumed that one-third of critical patient bed days would be treated in the general ward and two-thirds in the ICU.

The likelihood of additional COVID-related complications (per day) were estimated using evidence on the clinical course of COVID from patients in Wuhan, China, as were assumptions on the duration of symptoms (24,25). We assumed PCR tests would be used to determine COVID-19 status. The number of other diagnostic tests per hospitalisation was carried out in consultation with expert clinicians in essential critical care.

***Estimation of non bed-day costs***

An ingredients-based approach was used to calculate most of the service costs and prices for Pakistan. The data used was collected as part of the Disease Control Priorities 3-Universal Health Coverage (DCP3-UHC) project (15). Staff-related costs were constructed using federal-level pay scales. For most outputs, the number of minutes of staff required per activity were estimated via expert opinion obtained from clinicians working in the Health Planning, System Strengthening & Information Analysis Unit (HPSIU) in the Ministry of National Health Services Regulations and Coordination of Pakistan. For outputs where this was unavailable, health economists agreed a plausible assumed value.

Drug regimens were costed using resource use data obtained through expert opinion (HPSIU) and a number of price sources. An assessment of strengths and weaknesses of different price sources was conducted and hierarchy of sources was established. The primary source of price data was the Sindh Health Department Procurement Price list. If a price was unavailable, the Federal Wholesale Price List for Generic Medicines was used as a second option. As a last resort, private sector market prices were used.

Costs of supplies and equipment were similarly constructed. Resource use was determined through expert opinion (HPSIU) and price source hierarchy established. The primary source was the Medical Emergency Resilience Fund 2019-2020, and a secondary source was private sector market prices.

For additional diagnostic and radiology costs were estimated using available literature and market prices. We assessed strengths and weaknesses of different price sources. For example, we used the 'Costing and Pricing of Services in Private Hospitals of Lahore: Summary Report' as our primary source as it contained a methodological appendix that suggested that an ingredients-based approach consistent with ours was followed. If some prices were unavailable we used user fees from the Pakistan Institute of Medical Sciences, procurement prices from the Medical Emergency Resilience Fund procurement prices and user fees from the Aga Khan University Hospital. Space costs were estimated using data from budget documents from the Federal government (Islamabad Capital Territory Health Infrastructure PC-1).

***Estimation of bed-day costs***

We took an ingredients approach to estimating the costs of general ward and ICU ward bed days, as these were major cost drivers in our cost model. We estimated the plausible number of nursing hours per bed day in an LMIC setting through consultation with members of the research team who have expertise in critical care in LMICs. In ICU the assumption of nurse to patient ratio would be 1:1; in the general ward the ratio would be 1:6 during the day time and 1:20 in the night.

To understand the full range of inputs required we obtained the underlying costing data set provided by the authors of a recent primary costing of hospital-based care (26). The paper reports the results of a detailed activity-based costing in a hospital in Karachi, disaggregated by phase of care. We used the cost data for the ward stay phase, removing any supplies or equipment specific to the surgery, to estimate the average generic costs of a bed-day (27).

We estimated the additional costs of ICU beds compared to standard hospital beds using an ingredients-based approach to cost the equipment and supplies not present in standard hospital wards. We used the procurement price of equipment and assumed depreciation over ten (ventilators and suction pumps) or five years (all other equipment). Supply costs included central and arterial lines, ventilator tubing, and sedatives.

***COVID-19-specific costs: Personal protective equipment and hygiene***

We calculated the costs of personal protective equipment (PPE) per health worker per day (Table A3) and allocated a cost per PPE per minute to clinical staff. We also calculated costs of hygiene per bed day (Table A4). We estimated the costs of PPE and hygiene supplies using a list of necessary supplies from a COVID-related budget from the Ministry of Health of Pakistan, which included local prices sourced by the Aga Kahn University. This was complemented for other countries using the WHO's Essential Supplies Forecasting Tool (ESFT) (28). We divided supplies into single-use and disposable. We determined plausible quantities and useful life for supplies following clinical guidelines and expert opinion. See tables S4-5 below.

***Table S4. PPE costs per general ward bed day and per ICU bed day***

| Supply | Price (USD) | Useful life (days) | Quantity per day | Total per member of staff per day (USD) | Assumptions |
|---|---|---|---|---|---|
| *PPE for General Ward* | | | | | |
| *Single Use* | | | | | |
| Surgical Gowns | 0.20 | 1 | 1 | 0.20 | |
| Nitrile Gloves | 0.05 | 1 | 10 | 0.45 | |
| Latex Gloves | 0.04 | 1 | 10 | 0.39 | |
| Disposable Head | 0.03 | 1 | 4 | 0.10 | |
| Shoe Covers | 0.02 | 1 | 4 | 0.06 | |
| Surgical Masks | 0.08 | 1 | 10 | 0.77 | |
| *Reusable* | | | | | |
| Goggles | 11.61 | 90 | 1.5 | 0.19 | Assuming half a day for washing |
| Gum Boots | 19.35 | 90 | 1.5 | 0.32 | Assuming half a day for washing |
| **Total:** | | | | 2.50 | |
| *PPE for ICU* | | | | | |
| *Single Use* | | | | | |
| N-95 Masks | 0.84 | 1 | 4 | 3.35 | |
| Disposable apron | 0.20 | 1 | 1 | 0.20 | |
| Nitrile Gloves | 0.05 | 1 | 10 | 0.45 | |
| Latex Gloves | 0.04 | 1 | 10 | 0.39 | |
| Disposable Head | 0.03 | 1 | 4 | 0.10 | |
| Shoe Covers | 0.02 | 1 | 4 | 0.06 | |
| Surgical Masks | 0.08 | 1 | 10 | 0.77 | |
| *Reusable* | | | | | |
| Face Shields | 27.81 | 5 | 1.5 | 8.34 | Assuming half a day for washing |
| Goggles | 11.61 | 90 | 1.5 | 0.19 | Assuming half a day for washing |
| Gum Boots | 19.35 | 90 | 1.5 | 0.32 | Assuming half a day for washing |
| **Total:** | | | | 14.19 | |

***Table S5. Hygiene costs per general ward and ICU bed-day.***

| Supply | Price (USD) | Useful life (days) | Quantity per day | Total per ICU bed per day (USD) | Assumptions |
|---|---|---|---|---|---|
| *Single Use* | | | | | |
| Hand Sanitizers | 47.97 | 1 | 0.05 | 2.40 | 100ml use per day, price assumed to refer to bottle of 2000ml |
| Biohazard Bags | 0.23 | 1 | 1 | 0.23 | |
| Disposable bed sheets | 1.94 | 1 | 1 | 1.94 | |
| Disposable Tissue Boxes | 0.65 | 1 | 1 | 0.65 | 1 box per day, price assumed to refer to 1 box |
| Disposable Tissue rolls | 0.35 | 1 | 1 | 0.35 | 1 roll per day, price assumed to refer to 1 roll |
| Disinfectants (1L Dettol) | 3.23 | 1 | 0.25 | 0.81 | 250ml used per day, price refers to bottle of 1000ml |
| Liquid Soaps (250ml Dettol bottles) | 1.74 | 1 | 0.2 | 0.35 | 50ml used per day, price refers to bottle of 250ml |
| Ethanol (1L bottles) | 16.13 | 1 | 0.1 | 1.61 | 100ml used per day, price refers to bottle of 1000ml |
| Liquid Bleach | 2.58 | 1 | 0.25 | 0.65 | 250ml used per day, price assumed to refer to bottle of 1000ml |
| *Reusable* | | | | | |
| Waste Bins | 15.03 | 90 | 1 | 0.17 | |
| Mackintosh bed sheets | 9.68 | 90 | 1 | 0.11 | |
| Mops | 2.58 | 90 | 1 | 0.03 | |
| Dusters | 0.32 | 90 | 1 | 0.00 | |
| **Total:** | | | | 9.28 | |

***COVID-19- specific costs: Oxygen supplementation***

Oxygen supplementation therapy is the main form of treatment for COVID 19. There are different methods of oxygen delivery which utilise different types of supplies, equipment and require different average levels of oxygen flow. We calculated costs for 6 types of oxygen delivery techniques and assumed a distribution across severe and critical patients according to members of our research team with clinical expertise in critical care in LMICs (Table S6).

Oxygen therapy costs per bed-day were calculated by estimating the number of cylinders consumed in 24 hours at different flow rates, assumed to be 10L per minute in the general ward and 30L per minute in the ICU. Cylinder duration (hours) was estimated by dividing pressure by the number of litres per minute, assuming a standard cylinder size of 4.6kg, filled at 1,900 psi pressure (29). Cost per cylinder was obtained from the online catalogue of a multinational commercial manufacturer that is active in Pakistan (30).

***Table S6. Pathways of oxygen management***

| | | | "Real-world" scenario | | |
|---|---|---|---|---|---|
| | | | Severe case | Critical case | |
| | | | Severe pneumonia (15% of COVID cases) | Acute respiratory distress syndrome (5% of COVID cases) | |
| | | | General ward | General ward only | ICU |
| *Supplemental oxygen management type* | | | | | |
| % ventilator | | | 0% | 0% | 50% |
| % CPAP | | | 0% | 0% | 25% |
| % high-flow nasal cannula | | | 0% | 0% | 25% |
| % non-rebreather mask | | | 25% | 100% | 0% |
| % nasal cannula | | | 50% | 0% | 0% |
| % high-concentration mask | | | 25% | 0% | 0% |
| *Total % Patients in pathway* | | | 100% | 33% | 67% |

***Household costs of illness***

We estimated household costs of illness and care-seeking for COVID-19 using previously published data on the household-incurred costs associated with symptoms of tuberculosis (persistent cough and fever). We used a model previously used to pool household cost data from different settings to adjust costs for COVID-19 duration and age distribution (31). Direct costs to households included out-of-pocket payments for both medical and non-medical goods and services (32–34); these cost estimates were adjusted from South Africa to reflect Pakistan prices using GDP deflators and exchange rates from the World Bank (35). We estimated the likelihood of lost income due to sickness or caregiving by age using age-specific labour participation rates from the International Labour Organization (36). We also include a household cost of death, including funeral costs, and a loss of one year of income where the patient was an income earner (37).

### 8. Disability Adjusted Life Year Estimates

***Table S7. Disability Life Years per COVID-19 death***

| Scenario: | Base case | | Additional comorbidities | |
|---|---|---|---|---|
| **Age band** | **No discounting** | **3% discounting** | **No discounting** | **3% discounting** |
| 0-4 | 57.51 | 25.66 | 48.07 | 22.33 |
| 5-9 | 53.95 | 25.00 | 45.40 | 21.92 |
| 10-14 | 49.19 | 23.64 | 41.20 | 20.65 |

| | | | | |
|---|---|---|---|---|
| 15-19 | 44.40 | 22.06 | 36.95 | 19.17 |
| 20-24 | 40.19 | 20.85 | 33.24 | 18.02 |
| 25-29 | 36.20 | 19.68 | 29.73 | 16.90 |
| 30-34 | 32.25 | 18.37 | 26.25 | 15.65 |
| 35-39 | 28.43 | 16.98 | 22.91 | 14.32 |
| 40-44 | 24.81 | 15.57 | 19.75 | 12.99 |
| 45-49 | 21.29 | 14.03 | 16.71 | 11.54 |
| 50-54 | 17.92 | 12.37 | 13.81 | 10.00 |
| 55-59 | 14.75 | 10.65 | 11.13 | 8.42 |
| 60-64 | 11.88 | 8.93 | 8.74 | 6.88 |
| 65-69 | 9.43 | 7.35 | 6.74 | 5.49 |
| 70-74 | 7.53 | 6.11 | 5.25 | 4.45 |
| 75+ | 4.88 | 4.19 | 3.45 | 3.06 |

### 9. Impact of time-horizon on the Incremental Cost-Effectiveness Ratio (ICER)

The impact of vaccination on SARS-CoV-2 transmission dynamics will result in changes in the number of infections (also cases, hospitalisations and deaths) that may vary over time, and depend upon particular assumptions about vaccine and immunity characteristics. As a result, the estimated ICER can vary, and even oscillate up and down, as the analysis time-horizon is increased. To check the convergence of the ICER over the 10-year time-horizon of our main analysis we performed a sensitivity analysis using different different assumptions about costs, vaccine price, DALYs, vaccine campaign duration, and duration of natural immunity. The results are shown in supplementary figures 6 and 7.

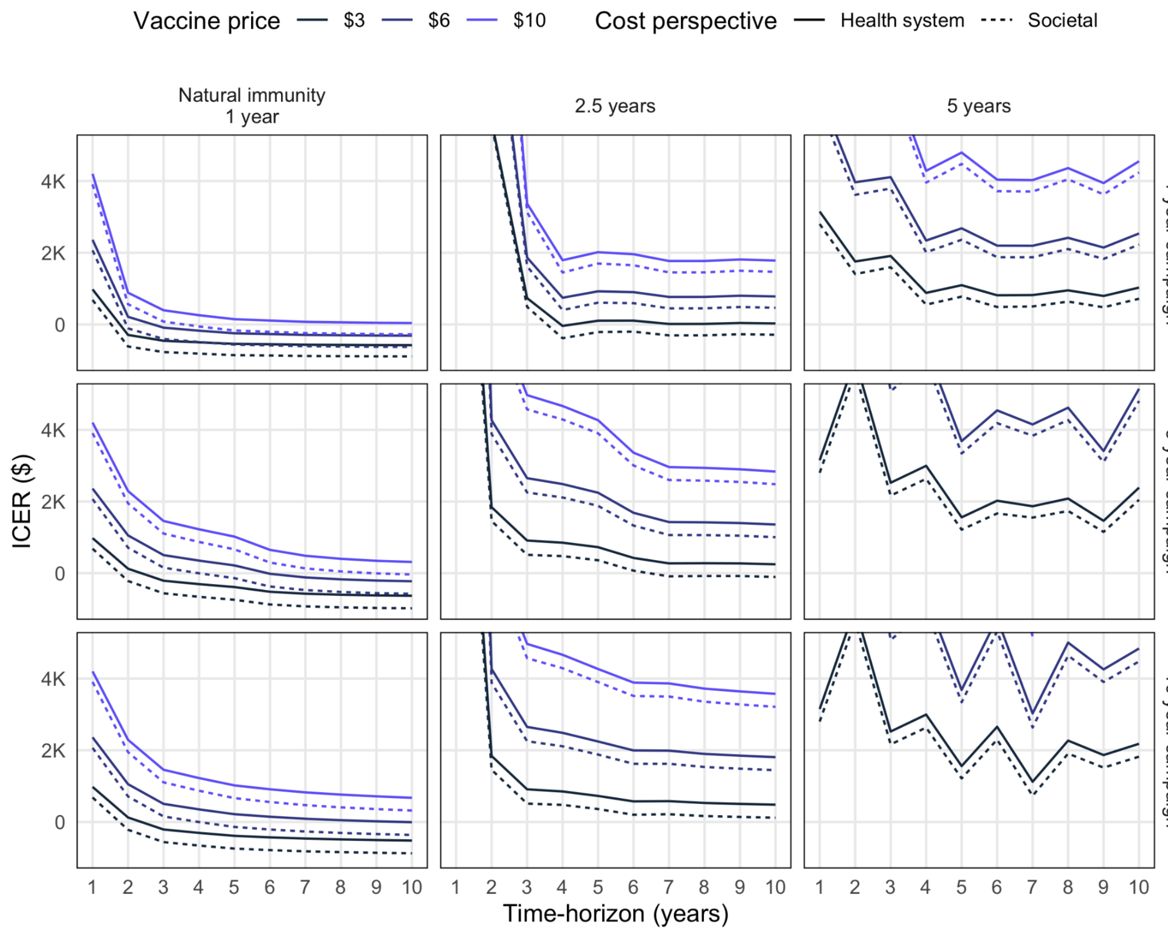


***Figure S6: Sensitivity of ICER to time-horizon under different assumptions about vaccine price, costs, campaign duration and duration of natural immunity.***

Results shown are for vaccination using a 2-dose regimen with 70% efficacy and 2.5y vaccine immunity duration, an initial vaccination rate of 4000 courses per day, and initial targeting of adults aged over 60 years old.

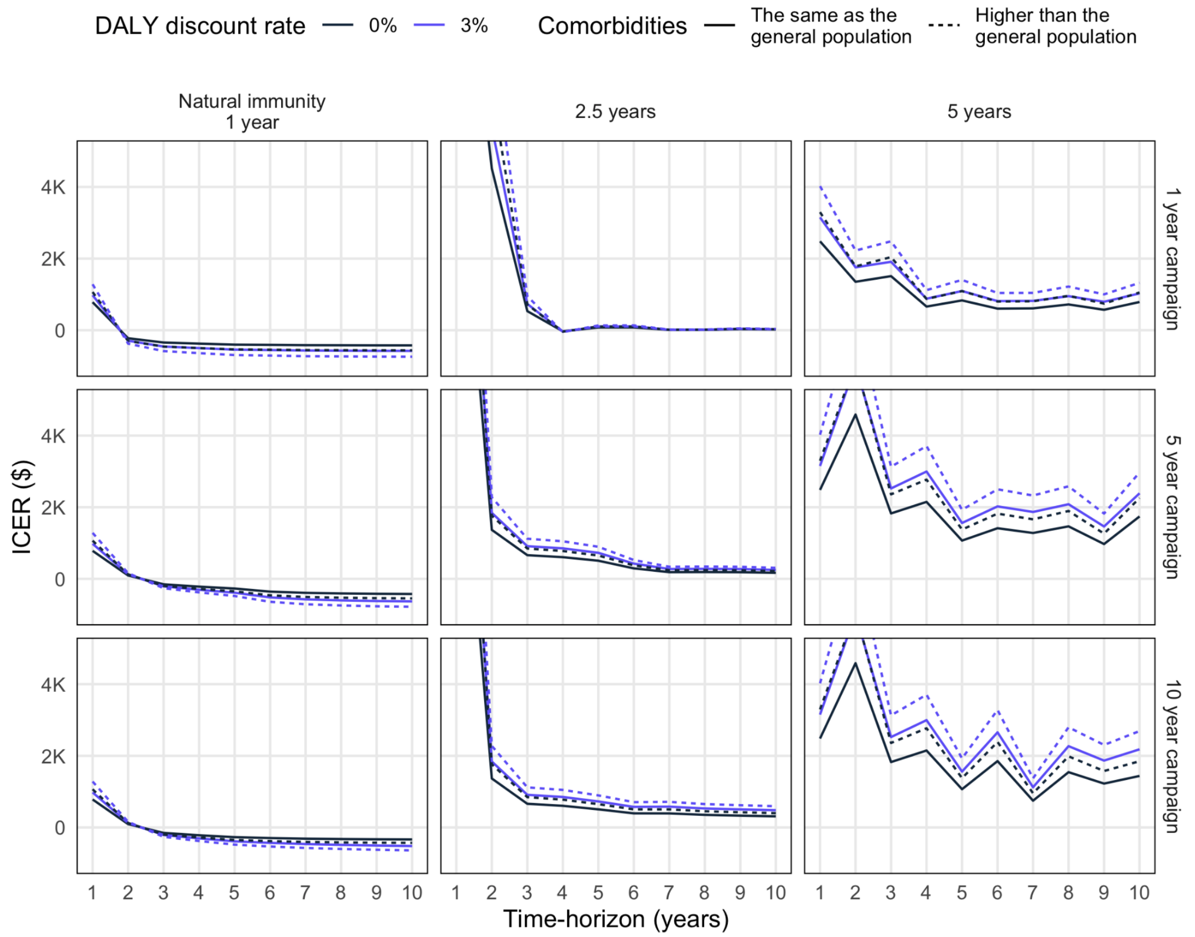


***Figure S7: Sensitivity of ICER to time-horizon under different assumptions about co-morbidities, discounting of DALYs, campaign duration and duration of natural immunity.***

Results shown are for vaccination using a 2-dose regimen with 70% efficacy and 2.5y vaccine immunity duration, an initial vaccination rate of 4000 courses per day, and initial targeting of adults aged over 60 years old.

### 10. Consolidated Health Economic Evaluation Reporting Standards (CHEERS) checklist

| Section/item | Item No | Recommendation | Reported on page No/ line No |
|---|---|---|---|
| Title and abstract | | | |
| Title | 1 | Identify the study as an economic evaluation or use more specific terms such as “cost-effectiveness analysis”, and describe the interventions compared. | See page 1. |
| Abstract | 2 | Provide a structured summary of objectives, perspective, setting, methods (including study design and inputs), results (including base case and uncertainty analyses), and conclusions. | The headings were adapted to fit the journal house style, but contain all the recommended items (see page 3):<br><br>Objectives: Mentioned in Background.<br><br>Perspective, setting, study design, inputs: Mentioned in Methods.<br><br>Base case and uncertainty: Mentioned in Results.<br><br>Conclusions: Mentioned in Interpretation. |
| Introduction | | | |
| Background and objectives | 3 | Provide an explicit statement of the broader context for the study. | The policy context is described in the Introduction (see paragraphs 1-5 in page 5). |
| | | Present the study question and its relevance for health policy or practice decisions. | Presented in the last paragraph of the Introduction on page 5. |
| Methods | | | |
| Target population and subgroups | 4 | Describe characteristics of the base case population and subgroups analysed, including why they were chosen. | The two target population scenarios (15+ and then 65+ followed by 15+) and their justification are stated in the last paragraph of the Methods subsection called “Vaccine Programme”. See the third full paragraph on page 8. |
| Setting and location | 5 | State relevant aspects of the system(s) in which the decision(s) need(s) to be made. | The setting and the decision makers are stated in the last paragraph of the Introduction (see page 5). |
| Study perspective | 6 | Describe the perspective of the study and relate this to the costs being evaluated. | The cost perspective is described in the first paragraph of the Methods subsection called “Costs” (see the first full paragraph in page 9). |

| | | | |
|---|---|---|---|
| Comparators | 7 | Describe the interventions or strategies being compared and state why they were chosen. | A vaccine campaign of 1 or more years, either focused on 65+ individuals initially or broadly targeted from the beginning. Various vaccine performance scenarios are considered. See pages 7-8 (Vaccine programme section), as well as SI pages 4-5. These were chosen to reflect planned COVAX facility distribution and available vaccine product performance. |
| Time horizon | 8 | State the time horizon(s) over which costs and consequences are being evaluated and say why appropriate. | The time horizon is described in the third paragraph of the Methods subsection called “Vaccine Programme”. See page 7. |
| Discount rate | 9 | Report the choice of discount rate(s) used for costs and outcomes and say why appropriate. | The discount rate is described and justified in the first paragraph of the Methods subsection called “Costs” (see the first full paragraph in page 9). |
| Choice of health outcomes | 10 | Describe what outcomes were used as the measure(s) of benefit in the evaluation and their relevance for the type of analysis performed. | Cases, deaths, Hospitalisation admissions and person-days, and DALYs averted; see page 8. |
| Measurement of effectiveness | 11a | Single study-based estimates: Describe fully the design features of the single effectiveness study and why the single study was a sufficient source of clinical effectiveness data. | NA |
| | 11b | Synthesis-based estimates: Describe fully the methods used for identification of included studies and synthesis of clinical effectiveness data. | Vaccine efficacy and duration assumptions are stated and justified in the third paragraph of the Methods subsection named “Vaccine programme” (see first full paragraph in page 8). |
| Measurement and valuation of preference based outcomes | 12 | If applicable, describe the population and methods used to elicit preferences for outcomes. | NA |

| | | | |
|---|---|---|---|
| Estimating resources and costs | 13a | Single study-based economic evaluation: Describe approaches used to estimate resource use associated with the alternative interventions. Describe primary or secondary research methods for valuing each resource item in terms of its unit cost. Describe any adjustments made to approximate to opportunity costs. | NA |
| | 13b | Model-based economic evaluation: Describe approaches and data sources used to estimate resource use associated with model health states. Describe primary or secondary research methods for valuing each resource item in terms of its unit cost. Describe any adjustments made to approximate to opportunity costs. | Resource use estimates, sources and valuation methods are summarised in Table 1 (see pages 9-10) and described more fully in the "Costs" subsection in Methods (see page 9). Further details are available in the Supplementary Information (pages 6-14). |
| Currency, price date, and conversion | 14 | Report the dates of the estimated resource quantities and unit costs. Describe methods for adjusting estimated unit costs to the year of reported costs if necessary. Describe methods for converting costs into a common currency base and the exchange rate. | See first paragraph of subsection entitled "Costs" in Methods (page 9). |
| Choice of model | 15 | Describe and give reasons for the specific type of decision-analytical model used. Providing a figure to show model structure is strongly recommended. | These are described in the Methods subsection named "Epidemiological model" (see paragraphs 1-2 in page 6). The model structure is shown in the first figure in the Supplemental Information (page 1). |
| Assumptions | 16 | Describe all structural or other assumptions underpinning the decision-analytical model. | These are described in the first two Methods named "Epidemiological model" (see page 6) and "Model fitting and projections" (see page 7), with parameterisation described in Table 1 (see pages 9-10). |

| | | | |
|---|---|---|---|
| Analytical methods | 17 | Describe all analytical methods supporting the evaluation. This could include methods for dealing with skewed, missing, or censored data; extrapolation methods; methods for pooling data; approaches to validate or make adjustments (such as half cycle corrections) to a model; and methods for handling population heterogeneity and uncertainty. | These are described in the first two Methods subsections titled “Epidemiological model” (see page 6) and “Model fitting and projections” (see page 7), with parameterisation described in Table 1 (see pages 9-10). |
| Results | | | |
| Study parameters | 18 | Report the values, ranges, references, and, if used, probability distributions for all parameters. Report reasons or sources for distributions used to represent uncertainty where appropriate. Providing a table to show the input values is strongly recommended. | This is shown in Table 1 (see pages 9-10). Distributions are given for all epidemiological parameters. Economic parameters are varied in scenario analyses rather than sampled from distributions. |
| Incremental costs and outcomes | 19 | For each intervention, report mean values for the main categories of estimated costs and outcomes of interest, as well as mean differences between the comparator groups. If applicable, report incremental cost-effectiveness ratios. | Incremental costs and cost-effectiveness ratios are given in Table 2 (see page 17). |
| Characterising uncertainty | 20a | Single study-based economic evaluation: Describe the effects of sampling uncertainty for the estimated incremental cost and incremental effectiveness parameters, together with the impact of methodological assumptions (such as discount rate, study perspective). | NA |

| | | | |
|---|---|---|---|
| | 20b | Model-based economic evaluation: Describe the effects on the results of uncertainty for all input parameters, and uncertainty related to the structure of the model and assumptions. | The result of varying epidemiological parameters is shown in the uncertainty ranges in Figures 1-3 (see pages 12-14). Other parameters are varied in scenario sensitivity analyses; the range of uncertainty is reflected in the scenarios in Figures 4-5 (see pages 15-16) and Table 2 (page 17). |
| Characterising heterogeneity | 21 | If applicable, report differences in costs, outcomes, or cost-effectiveness that can be explained by variations between subgroups of patients with different baseline characteristics or other observed variability in effects that are not reducible by more information. | Not applicable; the model did not examine particular subgroups. |
| Discussion | | | |
| Study findings, limitations, generalisability, and current knowledge | 22 | Summarise key study findings and describe how they support the conclusions reached. Discuss limitations and the generalisability of the findings and how the findings fit with current knowledge. | Key findings are summarised in the first two paragraphs of the Discussion (see page 18).<br><br>Limitations are discussed in paragraphs 4-5 of the Discussion (see pages 18-19). |
| Other | | | |
| Source of funding | 23 | Describe how the study was funded and the role of the funder in the identification, design, conduct, and reporting of the analysis. Describe other non-monetary sources of support. | The source of funding for study authors are listed at the end of the manuscript. The funders of this study had no role in the study design, data collection, data analysis, data interpretation, or writing of the manuscript. |
| Conflicts of interest | 24 | Describe any potential for conflict of interest of study contributors in accordance with journal policy. In the absence of a journal policy, we recommend authors comply with International Committee of Medical Journal Editors recommendations. | A declaration of interests statement is provided at the end of the text. |
